# Changes in pro inflammatory and regulatory immune responses during controlled human schistosome infection and the development of clinical symptoms

**DOI:** 10.1101/2023.09.01.23294933

**Authors:** Emma L. Houlder, Koen A. Stam, Jan Pieter R. Koopman, Marion H. König, Marijke C.C. Langenberg, Marie-Astrid Hoogerwerf, Paula Niewold, Friederike Sonnet, Jacqueline J. Janse, Miriam Casacuberta Partal, Jeroen C. Sijtsma, Laura H. M. de Bes-Roeleveld, Yvonne C.M. Kruize, Maria Yazdanbakhsh, Meta Roestenberg

## Abstract

Schistosomiasis is a prevalent helminthiasis, affecting over 230 million people worldwide, with varied, stage specific morbidity. Whilst the Th2 and regulatory immune responses in chronic infection have been relatively well studied, we have little understanding of human immune responses during acute infection. This is despite the initial infective stages being proposed as crucial targets for much-needed vaccine development. Here, we comprehensively map immune responses in male and female single-sex controlled human *Schistosoma mansoni* infection. Using unbiased, high dimensional techniques we show that human immune responses to male and female single-sex infection are comparable. An early Th1-biased inflammatory response was observed at week 4 post infection, which was particularly apparent in individuals experiencing symptoms of acute schistosomiasis. This included expansion of HLA-DR^+^ effector memory T cells, CD38^+^ monocytes and an increase in serum IFNγ. By week 8 post infection these inflammatory responses were followed by an expansion of Th2 and of regulatory cell subsets, including IL-10 producing CD4^-^CD8^-^ T cells, CD11c^+^ atypical memory B cells and serum IL-10. This study provides immunological insight into the clinical manifestations of acute schistosomiasis, as well as critical context through which to understand the development of immune responses observed in natural infection.

**One sentence summary:** Controlled human schistosome infection reveals cellular and cytokine responses to schistosome infection, with early inflammatory responses in symptomatic individuals at week 4 and a balanced Th1, Th2 and regulatory response in all participants by week 8.

## Introduction

Globally, over 230 million people are infected with schistosomes, mainly *Schistosoma mansoni (S. mansoni)* and *Schistosoma haematobium* (*1*). Schistosomes undergo multiple developmental stages in the human host, causing stage-specific morbidity. Cercarial dermatitis occurs in responses to skin invasion, followed by acute schistosomiasis syndrome (Katayama fever) 2-7 weeks later, in response to migrating and maturing schistosomes (*2–5*). Severe disease manifestations occur in up to 10% of infected individuals during patent (egg-producing) infection, with eggs released into the vasculature becoming lodged in multiple organs, inducing a granulomatous response and varied clinical presentations (*6*). Sterilizing immunity does not develop against schistosomes, meaning that post treatment with the schistosomocidal drug praziquantel individuals can be rapidly re-infected upon exposure to contaminated water (*7*). There is no approved vaccine for schistosomiasis. Moreover, vaccine design is hindered by a lack of understanding of natural immune responses to schistosomes, particularly during acute stages of infection, despite suggestions from pre-clinical models that larval antigens are key vaccine targets (*8*).

Our understanding of immune responses in acute human schistosomiasis has been guided by a limited number of clinical and immunological studies. Clinical manifestations of acute schistosomiasis include fever, fatigue, cough and eosinophilia (*2*). One endemic study, looking at individuals with acute schistosomiasis (4-8 weeks post exposure) has shown enhanced production of Th1 (T helper) cytokines (IL-1, TNFα, IFNγ) when compared to chronic schistosomiasis patients, who have higher levels of the Th2 associated cytokine IL-5 (*9*). In murine systems egg-production is the major driver of type-2 immune responses in schistosomiasis (*10*), with a mixed type-1/type-2 response present in pre-patent or single-sex *S. mansoni* infection (*11–13*).

Precise mapping of human immune responses over time has only become possible following the establishment of a controlled *S. mansoni* infection model (*3, 5*). In these studies, volunteers are infected with 10-30 single-sex cercariae, prior to treatment at week 8 and 12 (female cercariae) or week 12 (male cercariae) with praziquantel (*3, 5*). Acute schistosomiasis symptoms, including fever, headache and myalgia were observed in 14 of 30 participants between weeks 3 and 7 post infection, with 9 of these symptomatic participants experiencing a severe adverse event (*3, 5*). Similar immune responses were found in both studies with increased serum levels of the inflammatory chemokine CXCL10 at week 4, as well as increased schistosome specific Th1 (IFNγ) and Th2 (IL-4/IL-5/IL-13) cytokines (*3, 5*). Whilst informative, more detailed immune profiling is still required to understand cellular and cytokine changes during the initial months of schistosome infection.

Here, we used mass cytometry as an high-dimensional technique to profile immune responses in the first months of *S. mansoni* infection. Alterations in frequency and phenotype of peripheral blood mononuclear cells (PBMCs) were measured at weeks 4, 8 and 12 post infection with cellular results supported by multiplex serum cytokine analysis. Comparisons to baseline and between symptomatic and asymptomatic participants were performed in order to further our understanding of the pathogenesis of acute schistosomiasis syndrome.

## Results

### Immune cell composition and overall changes

To understand immune changes during schistosome infection, PBMC and serum samples were analyzed from two independent studies, male *S. mansoni* infection (n=14) at weeks 0, 4 and 8 post infection, and female *S. mansoni* infection (n=13) at weeks 0, 4, 8 and 12 post infection (Fig. 1A) (*3, 5*). Choice of timepoints was informed by clinical findings, with acute schistosomiasis symptoms occurring between weeks 3 and 7 post infection (Fig. 1A) (*3, 5*). As these were part of dose-escalating clinical safety trials, volunteers were experimentally exposed to 10 cercariae (n=6), 20 cercariae (n=10) or 30 cercariae (n=3) (*3, 5*). PBMCs were stained with a comprehensive panel encompassing 35 phenotyping markers, acquired, pre- processed and then clustered via self-organizing map (SOM) followed by hierarchical clustering (Fig. 1B). We were able to categorize 66 immune cell clusters into seven lineages - CD4^+^ T cells, CD8^+^ T cells, γδT cells, unconventional T cells, B cells, ILCs, and myeloid cells (fig. 1C, 1D).

**Figure 1.**
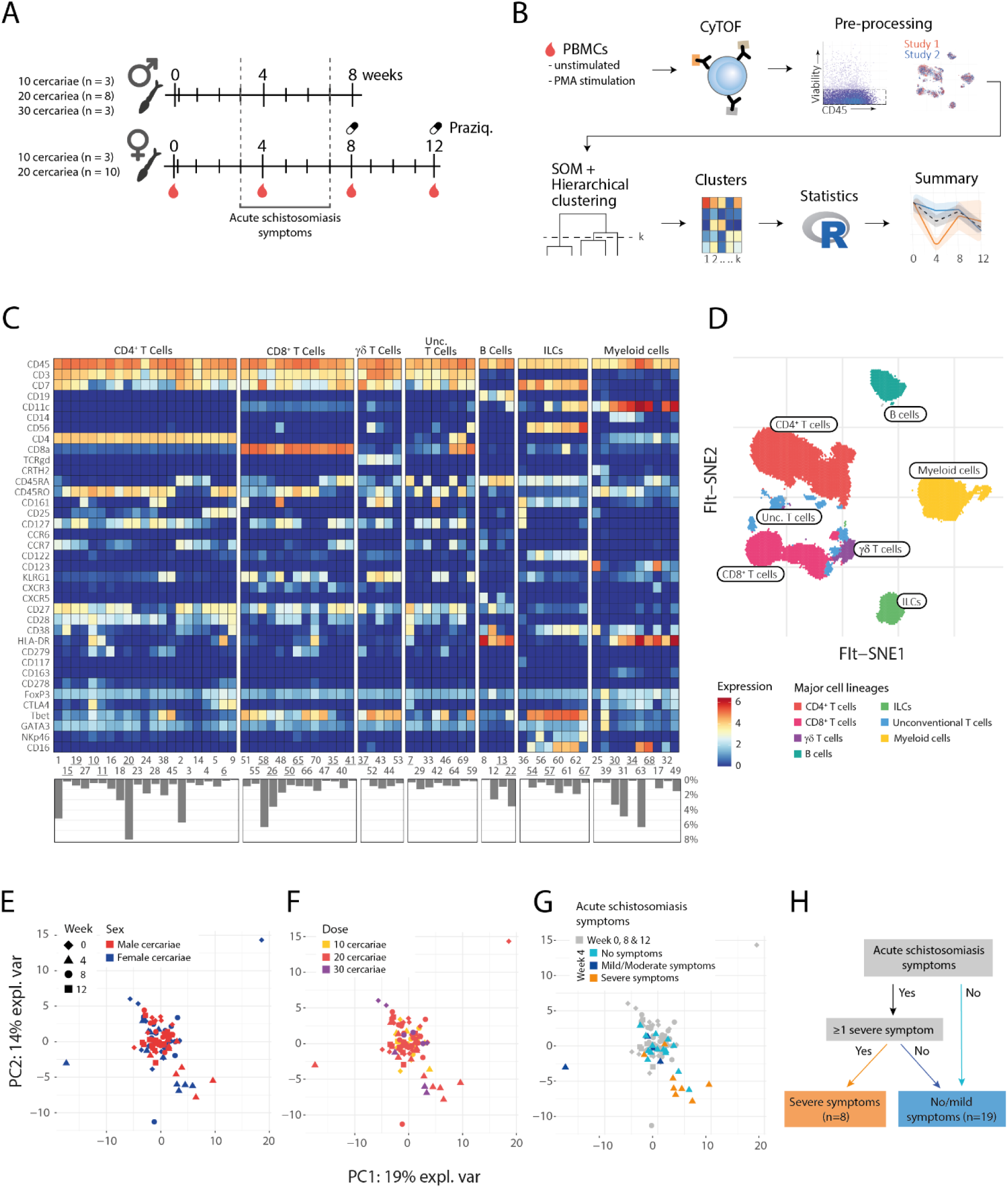
Overview of study design and cellular changes during controlled *S. mansoni* infection. **A**) Graphical representation of the trial design and **B)** analysis pipeline. **C)** Upper panel is a heatmap of identified cell clusters after CyTOF analysis of unstimulated samples, based on SOM and hierarchical clustering. Each tile depicts the median expression of a given marker (y-axis) for a specific cluster (x-axis). Lower panel depicts the abundance of each cluster. Percentage on the y-axis is the total cells of a given cluster divided by total CD45^+^ cells per sample. Clusters that were found significantly different from the linear mixed model are underscored for readability. **D)** Binned representation of the t-SNE map with CD45^+^ cells, colored by lineage of majority of cells in the bin. FIt-SNE is performed on a down sampled 4000 cells per sample. Bivariate binning is based on 150 xbins. **E-G)** A multilevel PCA was applied to individual samples, one sample being one participant at a specific timepoint, on the basis of cluster frequencies (scaled to unit variance). The PCA was colored based on **E)** sex, **F)** dose, or **G)** symptoms. **H)** Flow chart for classification of individuals into two groups; volunteers with severe symptoms or no/mild symptoms.

First - as an exploratory step, we performed principal component analysis (PCA) on the cluster frequencies, whereby we included all individuals on all timepoints. The first two principal components were able to explain 33% of the variation in the data (Fig. 1E). Notably, a group of 12 data points were separated from the rest on both axes. Coloring the data points by key clinical factors, including cercarial sex, dose and symptoms, allowed us to investigate their relative contributions to the principal components. Neither cercarial sex (Fig. 1E) nor dose (Fig. 1F) seemed to explain the largest block of variance. We formally tested for cercarial sex, with only two cell clusters changing differently over time between male and female infection (Supp. Fig. 1). These were cluster 25 (c25), basophils, which were increased at week 8 in male but not female infection and c49, CD14^-^CD16^-^HLA-DR^+^ conventional DCs, which increased at week 4 in female but not male infection (Supp. Fig. 1).

Next, we checked if symptoms explained the subset that were grouped separately in the PCA at week 4, shown in figure 1E-G as triangles. Interestingly, individuals who experienced only mild/moderate symptoms of acute schistosomiasis clustered with the asymptomatic group, whereas most individuals who experienced at least one severe symptom clustered separately (Fig. 1G). In contrast, individuals who had only mild or moderate graded acute schistosomiasis symptoms had immune responses that clustered with asymptomatic participants. For further analysis we have therefore grouped individuals with only mild or moderate symptoms with the asymptomatic participants, as explained in figure 1H. In text and figures these groups will be referred to as volunteers with either severe symptoms or no/mild symptoms. This categorization provides increased confidence in a correct diagnosis of acute schistosomiasis syndrome, which consists of a variety of aspecific symptoms including headache, fever, and malaise (*3, 5*).

### Inflammatory responses, enhanced in individuals with severe symptoms, observed at week 4 post infection

We next asked how immune responses changed during infection, taking into account whether individuals had experienced acute schistosomiasis symptoms. In not/mildly symptomatic individuals there were no visible differences between cell lineages over time (Fig. 2A). However, in individuals with severe symptoms at week 4 we can see clear changes in the overall immune composition. Myeloid cells are increased, while CD8 T and B cell lineages are decreased, with other timepoints remaining relatively stable compared to baseline (Fig. 2A).

**Figure 2.**
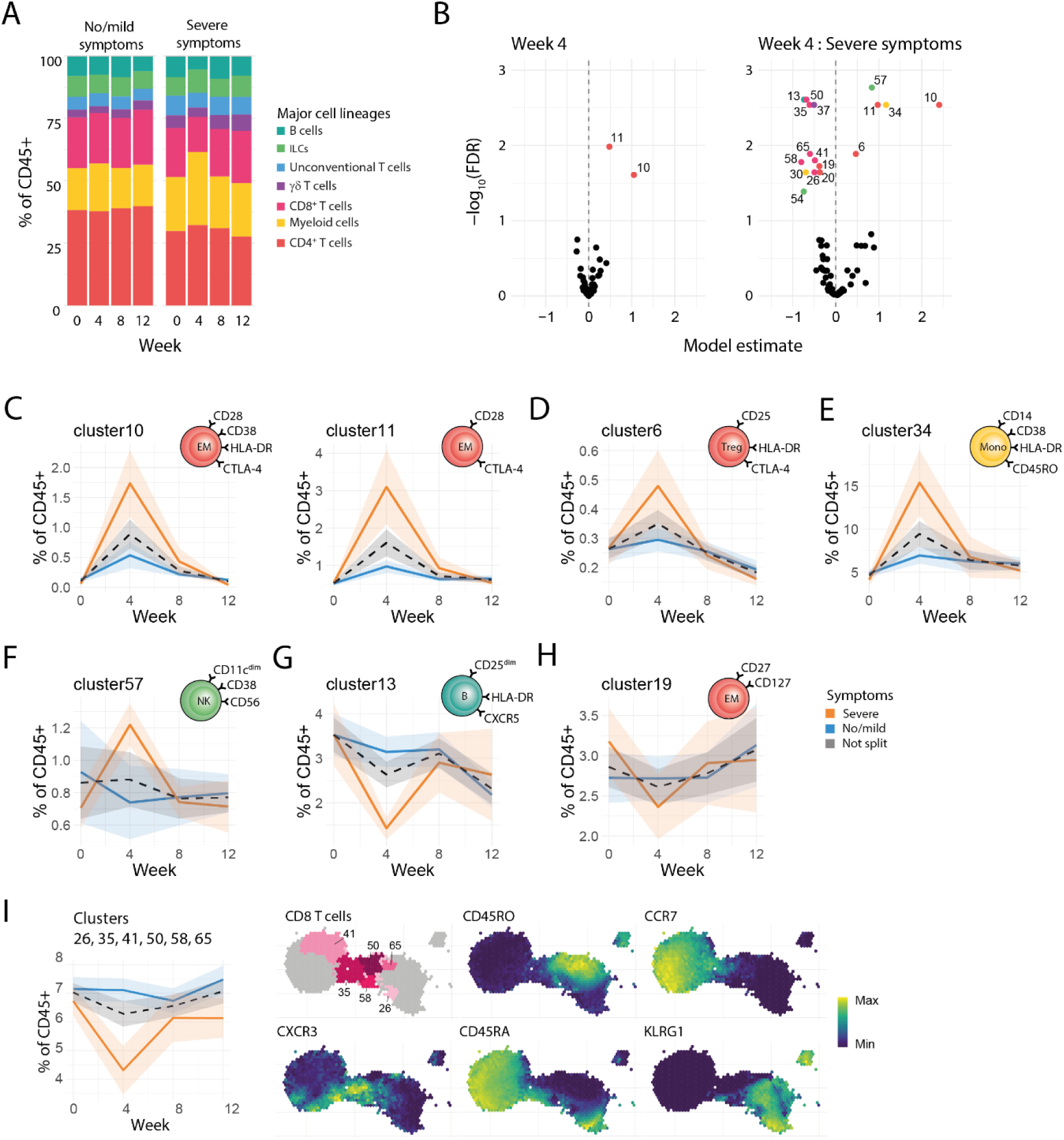
Characterization of cellular kinetics at week 4 of *S. mansoni* infection. **A)** Stacked bar chart of lineage frequencies in no/mildly symptomatic or severe symptomatic individuals. Each bar section represents the mean frequency of that cell lineage for the corresponding group. **B)** Volcano plots showing the model estimates and corresponding adjusted p values(-log10(fdr)) of the week 4 timepoint term (left), and the interaction term between week 4 and symptoms (right). Estimates are derived from a binomial linear mixed model, where each cluster is modelled separately. All analyses were performed on n=27 individual participants. Each point represents a cluster, clusters with fdr values < 0.05 are colored by lineage and labelled with cluster ID. **C-I)** Ribbon plots depicting mean (lines) frequency and standard error (shaded-area) of the mean of specified clusters. Cluster frequencies are given as percent of CD45^+^ cells and split/colored by symptom status. **I)** Right panel, binned representation of the CD8 cell island on the t-SNE map. Hex-bins colored by cluster or expression levels of selected markers. Clusters significantly altered at week 4 colored pink and non-altered clusters in grey. Left panel, Ribbon plots showing frequency of selected CD8 clusters (c50, c30, c65, c58, c41 and c26) over time.

To formally test changes in the immune cell composition we used a generalized linear mixed model to inquire how immune cell clusters changed over time, and whether these changes differed between symptom severity groups. Two CD4 T cell clusters, c10 and c11, increased significantly in all volunteers, however a significantly stronger increase in the abundance of these clusters was found for volunteers with severe symptoms (Fig. 2B) in comparison to volunteers with no/mild symptoms. In terms of phenotype c10 and c11 have similar immune markers, both activated HLA-DR^+^ effector memory (EM, CD45RA^-^CCR7^-^CD45RO^+^) CD4 T cells, differing in expression of CD38 (Fig. 1C, 2C). Notably, the expansion of these activated HLA-DR^+^ EM T cells was accompanied by an elevation of a HLA-DR^+^ CD4 Treg cluster (Foxp3^+^CD25^+^CD127^-^CTLA4^+^, c6), potentially providing early regulation of the inflammatory response (Fig. 2D) (*14*).

Innate inflammatory responses were also apparent in the severe symptomatic participants in contrast to the no/mildly symptomatic volunteers. Expansion of an activated CD38^+^ HLA-DR^high^ classical monocyte (CD14^+^CD16^-^) cluster (c34) was observed in severe symptomatic individuals (Fig 2E). Accompanying this was a decrease in a CD38^-^ HLA-DR^mid^ classical monocyte (CD14^+^CD16^-^) cluster (c30), suggestive of a transition towards more activation in the classical monocyte compartment in those developing severe symptoms (Fig. 2E, Supp. Fig. 4A & 5). PCA loadings (Supp. Fig. 2) reveal these myeloid and CD4 T cell clusters (c10, c11, c34 and c6) to be responsible for the observed separation of severe symptomatic week 4 samples in Fig. 1E-G. Finally, cytotoxic CD56^dim^ CD16^+^ NK cells (*15*), which tended to decrease in no/mildly symptomatic participants, significantly increased at week 4 in severe symptomatic individuals (Fig. 2F).

A number of lymphocyte clusters decreased at week 4 in symptomatic individuals. Potentially in response to chemokine signals, there was a reduction in CXCR5^+^CD38^-^ memory B cells (c13) at week 4 in individuals with severe symptoms (Fig. 2G). In addition, a modest reduction in two EM CD4 T cell clusters at week 4 was observed in severe symptomatic individuals (Fig. 2H & Supp Fig. 4F). Most notable was a decrease in multiple CD8 T cell clusters (c50, c30, c65, c58, c41 and c26) (Fig. 2I). To understand this reduction, we checked whether these clusters expressed similar phenotypic markers. The reduced CD8 T cell clusters had heterogenous expression of memory and naïve markers (CD45RO, CCR7, CD45RA), however all were KLRG1^+^ with the majority expressing CXCR3, which binds interferon inducible ligands including CXCL9 and CXCL10 (*16*). It is possible therefore, that the reduction in CD8 T cell clusters was in response to increases in CXCR3 ligands during the inflammatory response. Taken together cellular changes at week 4 are characterized by alterations in clusters from multiple cell lineages in symptomatic individuals, with expansion of mainly CD4 T cell clusters, monocyte activation, and decreases in chemokine receptor expressing CD8 T and B cells.

### Cellular responses at week 8-12 post infection

At week 8 post infection fewer significantly altered cell clusters were observed (Fig. 3A), with only one cluster elevated post treatment (week 12). Moreover, no significant interactions were observed between time and symptoms. This is in line with the subsiding of acute schistosomiasis symptoms by week 7 (*3, 5*), and indicative that the exaggerated inflammatory response observed in severe symptomatic individuals does not lead to altered long-term responses to the adult worm.

**Figure 3.**
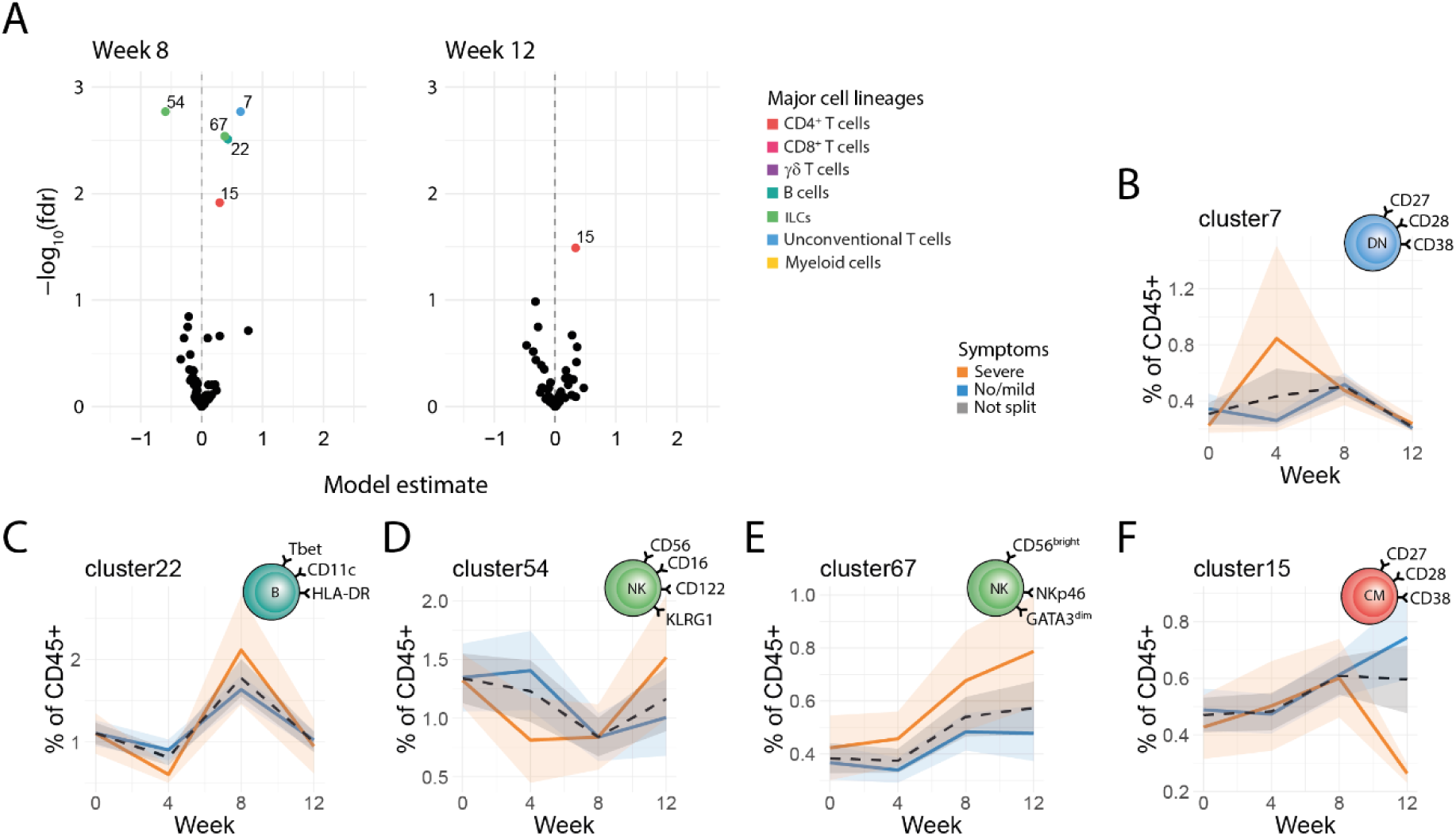
Characterization of cellular kinetics at week 8-12 of *S. mansoni* infection. **A)** Volcano plots showing the estimates and minus log scaled false discovery rate corrected p values (-log10(fdr)) of the week 8 timepoint term (left), and week 12 timepoint (right). Values derived from a binomial linear mixed model. Each point represents a cluster, clusters with fdr values < 0.05 are colored by lineage and labelled with cluster ID. **B-F)** All colored ribbon plots show of cluster frequencies as percent of CD45^+^ cells. These are split and colored by symptoms, with lines representing the mean value and ribbon shading ± standard error of the mean (SEM). **B)** Ribbon plots showing frequency of c7 over time. **C)** Ribbon plots showing frequency of c22 over time. **D)** Ribbon plots showing frequency of c54 over time. **E)** Ribbon plots showing frequency of c67 over time. **F)** Ribbon plots showing frequency of c15 over time. All analyses were performed on n=27 individual participants.

Cellular responses at week 8 tended to have a more mature or regulatory phenotype. For instance, there was an increase in CD38^+^ double negative (CD4^-^CD8^-^) T cells, c7 (Fig. 3B), which have previously been shown to inhibit Th1 responses (*17–19*). Additionally, an increase in atypical CD11c^+^Tbet^+^CD38^-^ atypical memory B cells (Fig. 3C) was observed, a B cell type typically expanded in chronic infections, aging or autoimmunity (*20, 21*). Finally, at week 8 post infection there was a significant decrease in a cluster of cytotoxic CD56^+^ CD16^+^ NK cells, and an increase in regulatory CD56^bright^CD16^-^ NK cells(Fig. 3E) (*22, 23*). The existence and dynamics of key clusters (c10/11, c34, c7 & c22) that changed at weeks 4 and 8 were independently confirmed by flow cytometry (Supp. Fig. 3E).

At week 12, post treatment, cellular responses tended to return to baseline. The one exception to this was a rare CD38^+^ central memory (CM, CD45RA^-^CCR7^+^CD45RO^+^) CD4 T cell cluster (c15), which increased at week 8 and remained elevated until week 12, perhaps representative of a sustained memory response (Fig. 3A & F).

### Cellular cytokines

Having established cellular dynamics during infection, we next investigated functional properties of these cells via assaying cytokine production post stimulation with PMA/ionomycin. First, we considered the global cytokine production as the total amount of cytokine positive cells divided by all CD45^+^ cells. At week 4 post infection the only significantly increased cytokine was Th2 (Il-13/IL-4/IL-5), which remained elevated up to week 8. At week 8 a mixed response was observed with decrease in the pro-inflammatory cytokine TNFα, as well as an increase in IL-6 and the regulatory cytokine IL-10 (Fig. 4A). No significant alteration in IFNγ, IL-2 or IL-17 expression was observed (Fig. 4A). To gain a more in-depth understanding, clustering was performed on stimulated cells, resulting in 70 clusters, 43 of which produced cytokine, the phenotype of these are summarized in Fig. 4B. To investigate, and formally test changes in frequency of cytokine-expressing cells, we used a similar binomial linear mixed model to inquire how immune cell clusters changed at week 4 and 8 post infection, and whether these changes differed in symptomatic individuals.

**Figure 4.**
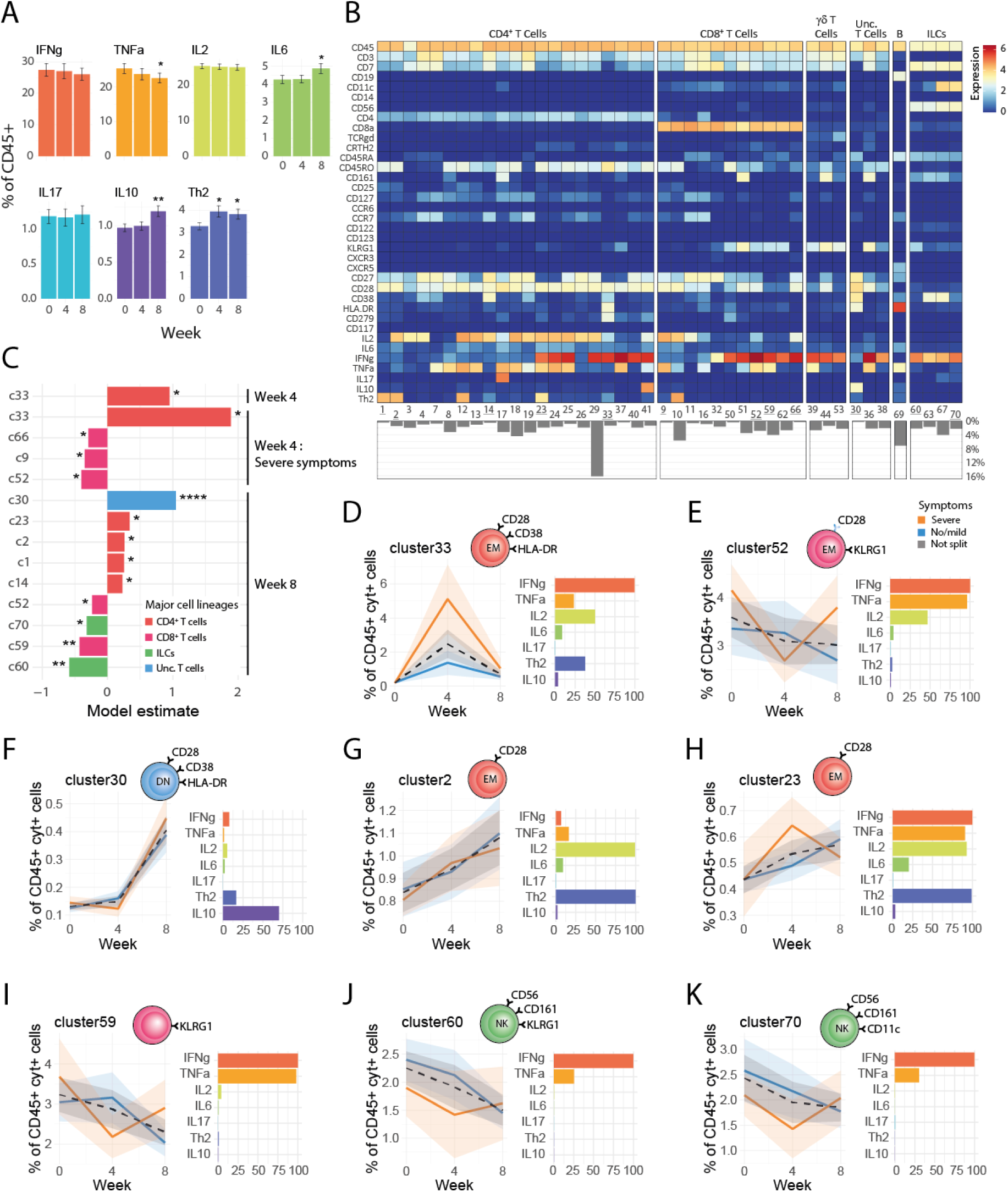
Characterization of cellular cytokine expression during *S. mansoni* infection. Cytokines were measured in PMA/ionomycin stimulated PBMC samples (n= 27 individual participants). **A)** Cytokine frequencies in total CD45^+^ cells. A binomial linear mixed model was used to determine changes from baseline. Error bars show the SEM. Uncorrected p values summarized in graphs as follows * p <0.05, ** p <0.01. **B)** Heatmap of identified cell clusters after CyTOF analysis of stimulated samples, based on SOM and hierarchical clustering. Only cytokine-expressing cell clusters are shown, with clusters in which less than 50% of the cells express any cytokine excluded. Each tile depicts the median expression of a given marker (y-axis) for a specific cluster (x-axis). **C)** Bar chart showing values derived from a binomial linear mixed model comparing cluster frequency over time and in relation to symptoms. Clusters with significant changes (fdr corrected p < 0.05) over time, with or without symptom interaction are shown. Bars are colored by lineage and labelled with cluster ID. **D-K)** All colored ribbon plots show cluster frequencies as percent of total CD45^+^ cytokine^+^ cells. These are split and colored by symptoms, with lines representing the mean value and ribbon shading ± standard error of the mean (SEM). Cytokine expression in each cluster is shown in the bar chart, colored by cytokine. FDR corrected p values summarized in graphs as follows * p <0.05, ** p <0.01, *** p <0.001, **** p < 0.0001

At week 4 post infection an increase in HLA-DR^+^ EM T cells, as well as a decrease in a number of CD8 T cell clusters was observed in individuals with severe symptoms, corresponding to our findings in the unstimulated panel. At week 4 there was an increase in c33, a HLA-DR+ EM T cell, which was elevated in severe symptomatic participants (Fig. 4D), phenotypically similar to the HLA-DR+ EM T cell we observed elevated in the unstimulated panel. These HLA-DR^+^ EM T cells were all IFNγ^+^, with a large minority also IL-2 and Th2 positive (Fig. 4D). Reduced CD8 T cell clusters (Fig. 4E and Supp. Fig. 6A&B) expressed Th1 cytokines IFNγ and TNFα. Taken together, Th1/Th2 cytokine expressing CD4 T cells increased, whereas Th1- cytokine expressing CD8 T cells decreased at week 4, with a higher magnitude of both effects in symptomatic individuals.

In contrast to our cellular findings, the majority of changes in cytokine-producing clusters were observed at week 8 post infection (Fig. 4C). Notably, as we saw in the unstimulated panel, cluster frequencies in severe symptomatic and no/mild symptomatic individuals converged at week 8 post infection, with no long-term effects of the earlier enhanced inflammatory response in severe symptomatic individuals. Specifically, we observed that CD38^+^ DN T cells, which we observed expanding in the unstimulated panel Fig. 3B, express the regulatory cytokine IL-10 (Fig. 4F, confirmed Supp. Fig. 3D), supporting our proposition that these cells play a regulatory role. A shift to a more Th2-dominant response was observed in the CD4 T cell compartment, with an increase in a number of CD4^+^ T cell clusters that expressed Th2 cytokines, and co-expressed IL-2, IFNγ and TNFα (Fig. 4 G-H and Supp. Fig. 6C&D). Decreases in a Th1 cytokine (IFNγ and TNFα) expressing CD8 T cell cluster and two NK cell clusters were observed (Fig. 4I-K). Taken together, the symptom-related Th1/Th2 CD4 T cell response at week 4 was replaced at week 8 by a symptom-independent increase in Th2-expressing CD4 T cell clusters and IL-10 expressing DN T cells, with a decrease in Th1 cytokine expressing CD8 T cells and NK cell clusters.

### Circulating cytokines

To complement our cellular findings, we performed a 96-plex immunoassay on serum cytokines and chemokines. In line with our cellular results, we observed the most changes in serum cytokines at week 4 post infection, with most proteins being only elevated in individuals with severe symptoms and not for asymptomatic volunteers, with the exception of IFNγ (Fig. 5A). In the individuals with severe symptoms, we saw elevation of IFNγ, interferon induced chemokines (CXCL11, CXCL9, CXCL11), other type-1 inflammatory mediators (TNF) and factors involved in cell survival, migration and differentiation (OPG, LIF, CDCP1) (Fig. 5A). The majority of the cytokines upregulated at week 4 remained significantly elevated at week 8 (Fig. 5B). In these individuals serum cytokines tended to decrease between week 4 and 8, whilst in no/mildly symptomatic individuals levels tended to increase or remain constant, leading to equivalent cytokine levels by week 8. In contrast to our cellular results, where increases in Th2 cytokines were observed at week 8, we did not observe any significant changes in type-2 cytokines measured (IL4, IL13, IL5). Notably, at week 8 post infection there was a clear elevation of the regulatory cytokine IL-10, which was also the only cytokine that remained significantly elevated at week 12 post infection compared to baseline (Fig. 5B).

**Figure 5.**
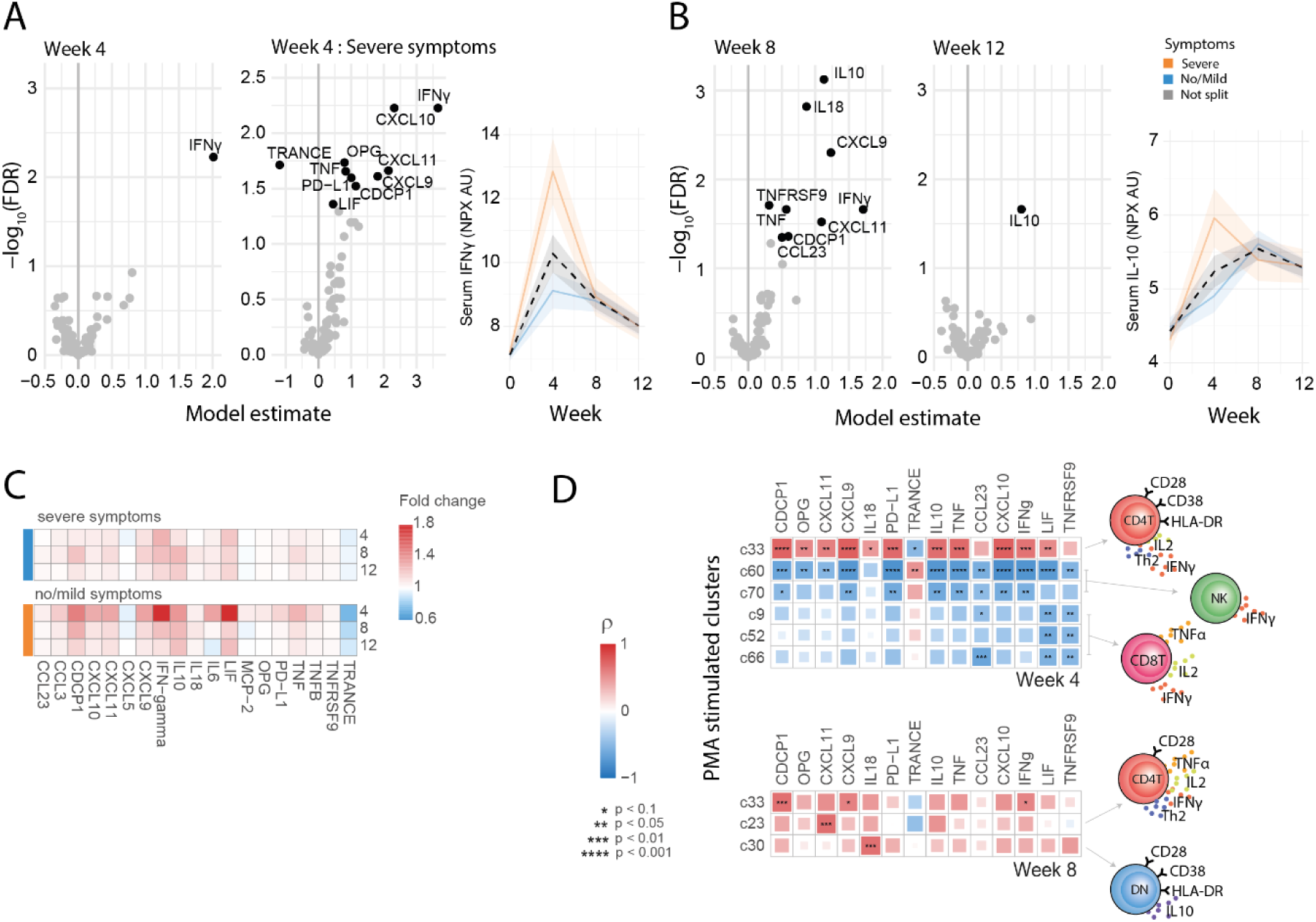
Characterization of serum cytokine kinetics during *S. mansoni* infection. A) Volcano plots showing the estimates and minus log scaled false discovery rate corrected p values (-log10(fdr)) of the week 4 timepoint term (left), and the interaction term between week 4 and symptoms (middle). Ribbon plots showing serum IFNγ over time (right). **B)** Volcano plots showing the estimates and minus log scaled false discovery rate corrected p values (-log10(fdr)) of the week 8 timepoint term (left), and the week 12 timepoint term (middle). Ribbon plots showing serum IL-10 over time (right). All values in volcano plots derived from a linear mixed model on NPX values (AU on log2 scale). Each point represents a cluster, clusters with fdr values < 0.05 are colored by lineage and labelled with cluster ID. Ribbon plots are split and colored by symptoms, with lines representing the mean value and ribbon shading ± SEM. **C)** Summary heatmaps showing mean fold change from week 0 of serum cytokines, split by symptomatic and asymptomatic individuals. Rows represent timepoint (weeks) and columns represent serum cytokines. **D)** Correlation matrices comparing frequencies of cell clusters (rows) and serum cytokines (columns) at weeks 4 and 8. Color and size of squares are proportional to the spearman’s p correlation coefficient. Cell clusters and serum cytokines selected to have significantly altered with time or symptoms, at a FDR<0.1. Significant correlations are shown, * =p<0.1, ** = p < 0.05, *** = p < 0.01, **** = p < 0.001

Correlation analysis was then performed to understand the interaction between serum cytokines and cellular cytokine responses at each timepoint (Fig. 5D). Stimulated cell clusters and serum cytokines were chosen for this analysis if they had been shown to significantly alter over time or having an interaction with acute schistosomiasis symptoms (Fig. 4C). Significant correlations were only observed at weeks 4 and 8 during schistosome infection, and not at baseline (Supp. Fig. 7). At week 4 IFNγ^+^ HLA-DR^+^ EM T cells (c33) were positively correlated with the majority of inflammatory serum cytokines, including IFNγ. At the same time, a negative correlation between in NK cell clusters c60 and c70 and the same inflammatory cytokines was seen (Fig. 4C,J&K). Together this suggests a concerted response regulating the observed decrease of NK cells and increase in IFNγ^+^ HLA-DR^+^ EM T cells at week 4 in symptomatic individuals. In contrast the decrease in CD8 clusters (c9,c52,c66) seen at this timepoint was negatively correlated to the cytokines CCL23, LIF and TNFRSF9, which have been proposed to have an inhibitory effect on CD8 T cells (Fig. 4C&I)(*24–26*). Fewer significant correlations were observed at week 8. Notably however serum IL-18 was significantly positively correlated to CD38^+^ DN T cells, and serum CXCL11 to Th2/IFNγ^+^/TNFα^+^ EM T cell c23, potentially suggestive of functional relationships between these cytokines and expansion of these cell clusters at week 8 post infection. Integrating cellular and serum cytokine responses has therefore provided insight into different networks that regulate the early inflammatory response at week 4, as well as the increasingly mature response at week 8.

## Discussion

This study has comprehensively delineated cellular and cytokine responses in the first months of schistosome infection, combining results from two single-sex (male or female) controlled human infection studies. Use of mass cytometry has allowed us to identify cell clusters across seven lymphoid and myeloid lineages in stimulated and unstimulated settings, complemented by multiplex immunoassay to assess serum cytokine responses. Inflammatory responses, including serum IFNγ and related pro-inflammatory cytokines, Th1/Th2 cytokine-producing HLA-DR^+^ EM T cells, and activated monocytes were elevated at week 4 in symptomatic individuals. By week 8 post infection responses in severe symptomatic and no/mildly symptomatic individuals had converged, with an expansion of regulatory IL-10^+^ DN T cells, atypical CD11c^+^ memory B cells as well as an increase in Th2 cytokine-producing CD4 T cells observed in all symptom groups. Post-treatment immune responses returned to baseline, with the exception of serum IL-10 and a CD4 CM T cell cluster which remained elevated.

At week 4, during the peak of reported symptoms, there were dramatic alterations in specific systemic immune cell clusters(*3, 5*). Whilst the exact timing of *S. mansoni* development in humans is unknown, rapid intra-mammalian growth (from 1 to 5mm) occurs between weeks 3 to 4 of infection, potentially accompanied by exposure of PAMPs to the immune system(*27, 28*). In reaction, we have observed expansion of activated CD38^+^ classical monocytes, particularly in symptomatic individuals. This monocyte phenotype has been previously reported in individuals that develop typhoid disease (including fever) following controlled human infection with the intracellular bacteria *S. typhi*, as well as during viral (COVID-19 and HIV) infection and active autoimmune response (systemic lupus erythematosus) (*29–31*). In addition, we observed expansion of Th1/Th2 cytokine-expressing HLA-DR^+^ EM CD4 T cells, which have also been shown to expand in other acute inflammatory conditions, including during active tuberculosis infection, where HLA-DR is a marker of recent cell division(*32*). The observed HLA-DR expression in CD4 T cells may derive from interaction with activated monocytes, with CD4 T cells able to acquire HLA-DR by trogocytosis, enhancing TCR signaling, survival and Th2 cytokine expression(*33, 34*). Our cellular findings were supported by serum inflammatory cytokines, with IFNγ-related cytokines in severe symptomatic individuals at week 4 post infection. These cytokines, including IFNγ, CXCL10 and TNFα, are elevated in many other acute inflammatory conditions and can directly induce symptoms such as fever (*35, 36*). Together, this suggests that the proinflammatory responses at week 4 may represent a stereotypical acute response to infectious agents, potentially induced by increased foreign schistosome antigen exposure, and responsible for acute schistosomiasis symptoms.

Later in infection, at week 8 responses in individuals who had experienced severe symptoms and not/mildly symptomatic individuals converge, with an increased Th2 response and a muted serum Th1 cytokines. Conventional dogma states that egg production is required for Th2 responses in schistosomiasis(*9, 10, 37*), however, this has recently been challenged by works demonstrating Th2 responses to immature worms in the first month of infection, prior to egg deposition (*11, 13, 38*). Our work furthers these findings, showing continued enhancement of Th2 responses up to week 8 post infection, in a single-sex egg-free system. Alongside Th2 cellular responses, an elevation of Th1 serum cytokines (IFNγ, IL-18, CXCL9) was observed when compared to baseline, although this was muted when compared to severe symptomatic individuals at week 4. The convergence of immune responses in severe symptomatic and not/mildly symptomatic individuals at week 8 could be due to increased immune regulation in symptomatic individuals, downregulating their response. Here, we found an early (week 4) increase in a Foxp3^+^ Treg cluster increased in these individuals, moreover our previous work has shown adult worm antigen-specific Foxp3^+^ Tregs at higher levels in symptomatic individuals at week 8 (*3, 5*).

Here, we have shown an enhanced regulatory response at week 8, including an increase in serum IL-10 levels. IL-10 has crucial regulatory and host-protective roles in schistosomiasis (*39–42*). Here we identified CD38^+^ DN T cells as key producers of IL-10 at week 8 post infection and revealed a novel correlation between this subset and levels of the pro-inflammatory cytokine IL-18 (*43*). DN T cells have not previously been studied in schistosomiasis, however several studies have linked DN T cells to a suppressive function in non-infectious diseases, including reducing the ability of CD4^+^ T cells to produce Th1 cytokines (*17–19*). There is also evidence for a regulatory shift in the B cell compartment at week 9, with an expansion of atypical memory CD11c^+^ B cells. This B cell subset is expanded in chronic exposure, including during parasite (malaria) infection, with anergic characteristics and diminished B cell receptor signaling (*20, 21*).

At the post treatment timepoint, week 12, immune responses tended back to baseline, with few long-term changes. Only the regulatory cytokine IL-10 in serum and a CD38^+^ CM T cell cluster (c15) remained elevated. This is in contrast to studies in endemic areas, which have observed elevated immune responses following praziquantel treatment(*44, 45*). For instance an increase in antigen-specific cytokine expression and reduction in Foxp3^+^ Tregs 6 weeks post *S. haematobium* treatment (*45*). The difference between our work and the endemic situation could be attributable to the acute nature of the controlled human infection, where schistosome-induced immune hypo-responsiveness has not fully developed(*46, 47*). Moreover, in the endemic situation re-infection as well as schistosome egg persistence will, provide continued schistosome antigen stimulation post-treatment, which will not be present in this single-sex model. Responses at week 12 may also be underestimated due to the lower sample size at this timepoint, with only female-infection samples (n=13) measured, reducing statistical power.

Throughout infection we have shown only minor differences in immune response between female and male single-sex infection with schistosomes. This finding is in line with the initial immunological analysis we have performed on the separate controlled human infection studies(*3, 5*), as well as a recent murine study which found no immunological differences in chronic male or female single-sex infection(*48*). However, it is still surprising in the context of prior papers that have shown sex-related differences in immune priming, as well as the known morphological and transcriptional differences between male and female worms (*49–52*).

The combination of the controlled human infection model and high dimensional analysis methodology provides a unique opportuntiy for assessment of immune cell changes in human schistosome infection. This paper has particular relevance for understanding the pathogenesis of acute schistosomiasis in travelers to endemic countries (*53*). Specifically, we propose that Th1/Th2 cytokine-expressing HLA-DR^+^ EM CD4 T cells, CD38^+^ monocytes and pro-inflammatory serum cytokines are critical in driving acute schistosomiasis symptoms. Perhaps suprisingly, enhanced responses in severe symptomatic individuals at week 4 did not lead to enhanced immune responses at week 8. Instead, a compensatory regulatory and Th2 response appeared independent of symptoms, with expansion of IL-10^+^ CD38^+^ DN T cells and atypical memory CD11c^+^ B cells. The week 8 response aligns best with our understanding of responses in endemic infection, where acute symptoms are rare (*54, 55*). Notably in our work Th2 and regulatory responses developed in response to adult worms, contrasting the dogma that the development of these mature responses requires egg production(*56*). Using a high dimensional and semi-unsupervised approach we have highlighted unforseen cellular and cytokine changes that occur during the initial stages of schistosome infection. We do not know whether the immune changes highlighted here are protective against infection or symptoms in schistosomiasis. However, by highlighting the key players in the initial immune response in schistosomiasis, this work provides direction for future studies to unpick what constitutes a protective immune response, and therefore to guide schistosome vaccine development.

## Materials and methods

### Study design

Samples in this study were part of two dose-escalating clinical safety trials of controlled human schistosome infection. In the first study (NCT02755324) 17 volunteers were infected with 10 (n=3), 20 (n=11) or 30 (n=3) male *S. mansoni* cercariae, as previously described(*3*). Here we analyzed samples from 14 volunteers, exposed to 10 cercariae (n=3), 20 cercariae (n=8) or 30 cercariae (n=3). In the second study (NCT04269915) 13 volunteers were infected with 10 (n=3) or 20 (n=10) female *S. mansoni* cercariae, as previously described(*5*). Samples from all volunteers in this female infection study were analyzed. Volunteers were treated with praziquantel (40 mg/kg) at week 12 in the male infection study and at weeks 8 and 12 (60 mg/kg) in the female infection study. Both studies were carried out in the LUMC, Leiden from 2019-2022. The study was approved by the LUMC Institutional Medical Ethical Research Committee (Institutional Review Board P16.111 and P20.015). It was performed according to the European Clinical Trial Directive 2001/20/EC, in accordance with ICH-GCP guidelines and the Declaration of Helsinki. Written informed consent was obtained from all participants.

### PBMC cryopreservation and thawing

Heparinized venous blood was diluted 2x in Hanks’ balanced salt solution (HBSS, ThermoFisher) containing penicillin G sodium (100 U/ml, Euroco-pharma BV) and streptomycin (100 µg/ml, Sigma). Density separation was performed, with 10mL of Ficoll (Apotheek LUMC), added below diluted blood and samples centrifuged (400xg, 25min, low brake). PBMCs were collected, washed and cryopreserved in freezing media: 20% of heat-inactivated fetal calf serum (FCS; Bodinco), 10% dimethyl sulfoxide (DMSO, Millipore) in complete RPMI (RPMI 1640 (Invitrogen) containing pyruvate (1 Mm, Sigma), l-glutamine (2 mM, Sigma), penicillin G sodium (100 U/ml, Euroco-pharma BV) and streptomycin (100 µg/ml, Sigma)). Cells in freezing media were placed in a Nalgene Mr. Frosty Freezing Container (ThermoFisher), overnight at −80°C, prior to liquid nitrogen storage. PBMC samples were cryopreserved within 6 hours post blood collection.

### Mass cytometry

Prior to mass cytometry staining cryopreserved PBMCs were thawed at 37°C in 50% FCS/complete RPMI, washed twice in 10% FCS/complete RPMI and then placed on ice. 3 million cells were transferred to 5ml microcentrifuge tubes (Eppendorf) for direct non-stimulated staining or 5ml round-bottom falcon tubes (BD Biosciences) for PMA/ionomycin stimulation. Stimulated cells were incubated for 6 hours at 37°C with PMA (100 ng/ml; Sigma) and ionomycin (1 µg/ml; Sigma), with brefeldin A (10 µg/ml; Sigma) added for the final 4 hours. Stimulated and non-stimulated cells were then washed twice in Maxpar staining buffer (Fluidigm). All washes during extracellular staining were performed at 400 xg for 5-7 minutes. For the male infection study a barcode mix targeting β2 microglobulin (B2M) and CD298 was added to each individual sample in a 6-choose-2 scheme using palladiums 104, 106, 108, 110 and platinums 194 and 198 for 30 minutes at room temperature. For the female infection study a barcode mix targeting β2 microglobulin (B2M) was added to each individual sample in a 6- choose-3 scheme using cadmiums 106, 110, 111, 112, 114 and 116 for 30 minutes at room temperature. Cells were washed twice in staining buffer, and combined into batches of 6 to 20 samples, including one reference per batch. Batched cells were incubated in 1 ml of 500x diluted 500 µM Cell-ID Intercalator-103Rh (Fluidigm) for 15 minutes at room temperature. Cells were then washed in staining buffer, resuspended in staining buffer with 5% Human TruStain FcX Fc-receptor blocking solution (BioLegend) added for 10 minutes at room temperature. <u>M</u>etal conjugated antibodies for extracellular antigens (detailed in Supp. Table 1&2) were then added for 45 minutes at room temperature. Cells were washed twice with staining buffer, and from this point stimulated and non-stimulated cells were treated differently.

For the non-stimulated panel in the female infection study cells were fixed in 1.6% formaldehyde (Pierce) for 10 minutes at RT prior to permeabilization. In the male study this step was skipped and cells went straight to permeabilization with freshly prepared Fix/Perm (eBioscience Foxp3/Transcription Factor Staining Buffer set, prepared to manufacturer’s instructions) for 30-45 minutes at 4°C. Cells were then washed twice in Perm buffer (eBioscience Foxp3/Transcription Factor Staining Buffer set, prepared to manufacturer’s instructions). This, and all subsequent washes were performed at 800 xg for 5-7 minutes. Metal conjugated antibodies for intranuclear antigens (detailed in Supp. Table 1) were then added in perm buffer for 30 minutes at room temperature.

For the stimulated panel in the female infection study cells were fixed in 4% formaldehyde (Pierce) for 10 minutes at RT prior to permeabilisation. In the male infection study this step was skipped and the cells went straight to permeabilization with MaxPar Fix I Buffer (Fluidigm, prepared to manufacturer’s instructions) for 20 minutes at room temperature. Cells were then washed twice with MaxPar Perm-S Buffer (Fluidigm, prepared to manufacturer’s instructions) prior to addition of metal conjugated antibodies (detailed in Supp. Table 2) for intracellular staining for 30 minutes at room temperature. After intracellular staining, cells were washed twice with staining buffer.

After staining, stimulated and non-stimulated cells were fixed with 1.6-4% formaldehyde (Pierce) for 10 minutes at room temperature. Cells were washed and 125 µM Cell-ID Intercalator-Ir (Fluidigm) added in MaxPar Fix and Perm buffer (Fluidigm) overnight at 4°C. The following day cells were washed twice in staining buffer prior to cryopreservation as performed on fresh samples.

All timepoints from one participant were stained and measured together in a total of 11 batches, 4 in the male infection study and 7 in the female infection study. Inclusion of a reference PBMC in each batch allowed for quality control, ensuring comparability between batches. Samples were measured with a Helios CyTOF mass cytometer (Fluidigm) with Wide Bore, tuned according to Fluidigm’s recommendations. Prior to acquisition, cells were thawed in RPMI 50% FBS, washed and counted. Cells were resuspended in Cell Acquisition Solution (CAS, Fluidigm) in the male study, and water in the female study containing 10% EQ Beads (Fluidigm) for acquisition. Next to channels used to detect antibodies, channels for intercalators (103Rh, 191 and 193 Ir), calibration beads (140Ce, 151Eu, 153Eu, 165Ho 175Lu) and background/contamination (131Xe, 133Cs, 138Ba, 206Pb, 208Pb) were acquired. FCS files were normalized and concatenated in Helios software, without removing beads.

### Mass cytometry data analysis

After normalization the FCS files were auto gated on the gaussian parameters using CyTOFClean (v10.3(*57*)). Subsequently, gates were set to select single, live, CD45^+^ cells for each batch, using the openCyto package in R (Supp. Fig. 8). In the last two steps, the batch FCS files were compensated and de-barcoded using the CATALYST package in R (v1.16.2(*58*)). First, the compensation was performed using the non-negative linear least squared method, provided with a self-measured spillover matrix. Lastly, the single FCS files per sample were acquired by the default de-barcoding pipeline provided by CATALYST.

To account for variation between batches stained and acquired at different times batch correction was performed using the CytoNorm package(v 1.16.2 (*59*)).This approach first clusters the data prior and performs batch correction within each cluster, with k=10 chosen for the non-stimulated panel and k=5 chosen for the PMA/ionomycin stimulated panel. For each approach reference samples used for correction were produced by randomly sampling 200000 cells from each batch. Quality of the batch correction was assessed and representative tSNE plots pre and post batch correction are shown in Supplementary Figure 9.

To explore the mass cytometry data produced samples were clustered and visualized in a two dimensional tSNE map. Clustering was performed based on expression of all surface and intracellular antigens using a two-step method. First, cell expression was mapped onto a 15x15 SOM-grid using the Kohonen package (v3.0.11(*60*)). The SOM grid was then divided into meta-clusters using hierarchical clustering, with k=70 clusters chosen for the unstimulated panel and stimulated panel. Clusters were manually assigned to cell lineages based upon expression of well-defined lineage markers. Heatmaps of cluster marker expression were produced using the package cytofast (v1.3.3(*61*)). To understand the overall structure of the data cells were placed on a tSNE map using the FIT-SNE algorithm (v1.2.1(*62*)). For visualization this map was then binned using the hexbin package (v3.0.11(*63*)) with 150 bins allowing assessment of cell lineages, clusters or marker expression.

For the PCA analysis, cell frequencies were scaled to have unit variance. To address the repeated measurement design, we have applied the multilevel approach implemented in the Mixomics package (*64*). In this fashion we tried to minimize the variance between volunteers and focused on variation introduced by clinical outcomes.

### Serum cytokine analyses

Serum cytokine analysis was performed using the Olink platform. Serum samples were run at Olink in Uppsala (Sweden) or UMC Utrecht (Netherlands) using the Olink Target 96 inflammation panel covering 92 different proteins. Data are reported as NPX, an Arbitrary Unit (AU) on a log2 scale. Values below the limit of detection (LOD) were extrapolated and included in analysis. The majority of proteins (66 of 92) had all values above the LOD, with 16 proteins detectable in over 50% of samples (IL6, FGF-21, IL-15RA, MCP-3, NT-3, FGF-5, ST1A1, SIRT2, NRTN, IL-2RB, TSLP, IL-24, GDNF, ARTN, IL4, IL-20) and 10 proteins detectable in less than 50% of samples (IL13, IL-20RA, IL-22 RA1, IL5, LIF, FGF-23, IL-1 alpha, IL2, IL33, Beta-NGF).

### Statistical analyses

A similar modelling strategy was employed for the different modalities of data we’ve acquired. For the cellular analysis we used the cluster frequencies as primary outcome and for the serum cytokine data each single target was considered. In this univariate fashion, a generalized linear mixed model was employed with sex (of volunteer), timepoint (in weeks, as factor) and symptoms (dichotomous) as explanatory variable. As random effects both volunteer and study were added as random intercept. To match the nature of the data, a gaussian family was considered for the serum cytokine data and a binomial family with logit link for the cellular data. The frequency and weights for the binomial model were based on the CD45+ cells for the unstimulated data and for the PMA stimulated data CD45+ and cytokine positive cells. To deal with any under- overdispersion an extra random intercept for sample ID was added to the models for the cellular data. From the generated models the estimates together with their p-values (t-statistic, via Satterthwaite’s degrees of freedom method) were extracted. Afterwards for each modality of data, all p-values from the generated models were corrected with the Benjamini-Hochberg procedure. Adjusted p-values with FDR < 0.05 were considered significant. Analysis was performed in R (v4.2.1(*65*)) with the lme4 (v1.1(*66*)) and lmerTest (v3.1(*67*)) packages.

The correlation analysis between the stimulated cellular data and the serum data were performed on only a significant (fdr < 0.5) subset of clusters and proteins. For each week containing data from both studies (week 0, 4, 8) a correlation matrix was constructed based on spearman’s correlation. A significant association deviating from zero was tested for each pair of cluster and protein. For each week the resulting p-values were corrected with Benjamini-Hochberg procedure.

## Data Availability

All data produced in the present study are available upon reasonable request to the authors.

## Acknowledgements

MY, MR, KS and EH were responsible for study design. KA and EH were responsible for statistical analysis, data interpretation and prepared the first draft. MY, MR and EH acquired funding. M.A-H, JK, MC and MR were the clinical investigators. JJ performed trial management. MC-P, JS and JJ were responsible for production and release of cercariae. MK, EH, PN, FS generated the data and optimised the experimental protocols. LdB and YK co-ordinated PBMC sample collection. All authors reviewed the manuscript.

We thank P. van Genderen, A. Bustinduy and A. Wajja for their valuable advice as members of the safety monitoring committee. We thank O.A.C. Lamers, C.de Dood, S.T. Hilt, A. Ozir- Fazalalikhan, V.P. Kuiper, G.V.T. Roozen, L. Wammes, L. van Lieshout, G.J. van Dam,I.M. Amerongen-Westra, P. Meij, P.L.A.M. Corstjens, S. P. Jochems, A. van Diepen, C.H. Hokke, M.A.A. Erkens, J.L. Fehrmann-Naumann, M.S. Ganesh, H. Gerritsma, G.C. Hardeman, P.T. Hoekstra-Mevius, Y.D. Mouwenda, H.H. Smits, K. Suijk-Benschop, J.J.C. de Vries and C.J.G. van Zeijl-van der Ham for their laboratory, clinical and data-analyzing support during the study. Most of all we thank all volunteers participating in the study, without whom the study could not have been performed. This project has received funding from the European Union’s Horizon 2020 research and innovation programme under grant agreement No 815643.

Data and material availability: data will be available from the authors upon reasonable request.

