## Supplementary figures for "Changes in pro inflammatory and regulatory immune responses during controlled human schistosome infection and the development of clinical symptoms"

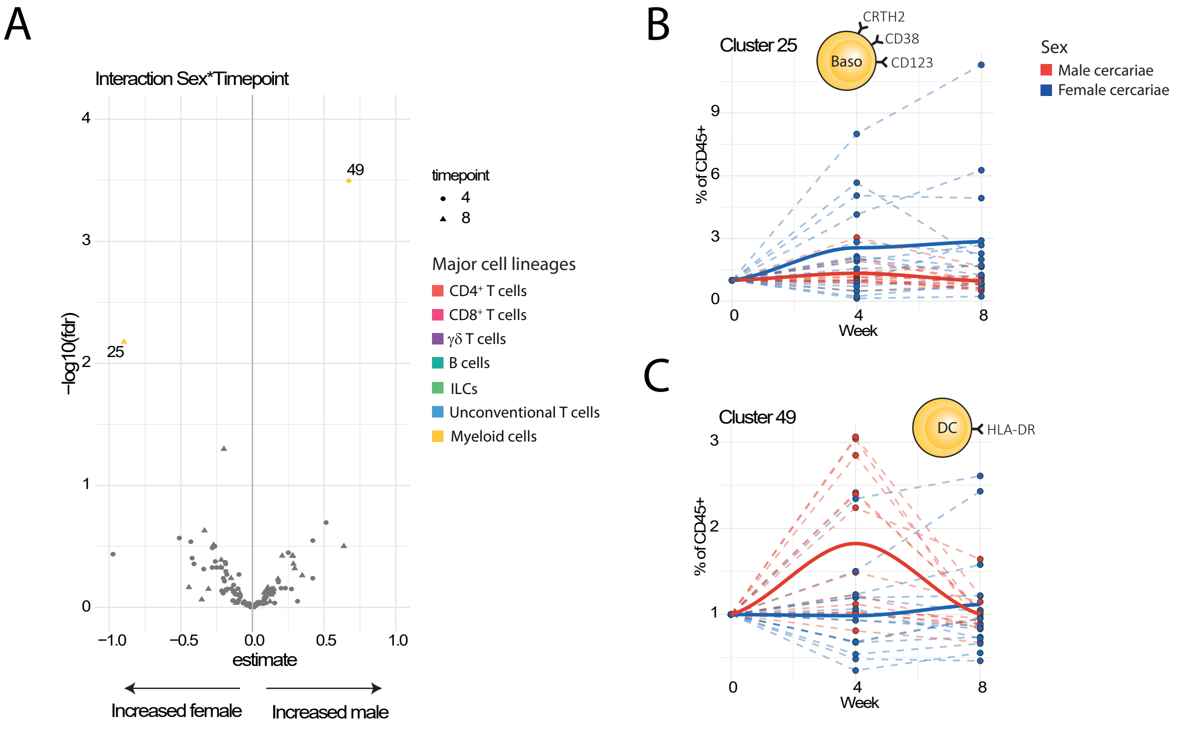


**Supplementary figure 1: Altered cellular dynamics in male and female cercariae infection.** A) Volcano plots showing the model estimates and corresponding adjusted p values(-log10(fdr)) of the interaction term between week 4 and 8 timepoint and cercarial sex. Estimates are derived from a binomial linear mixed model, where each cluster is modelled separately in a univariate manner. Each point represents a cluster, clusters with fdr values < 0.05 are colored by lineage and labelled with cluster ID. B) Line plot depicting frequency per participant (dashed) and mean (bold) of c25 over time, colored and split by cercarial sex. C) Line plot depicting frequency per participant (dashed) and mean (bold) of c49 over time, colored and split by cercarial sex.

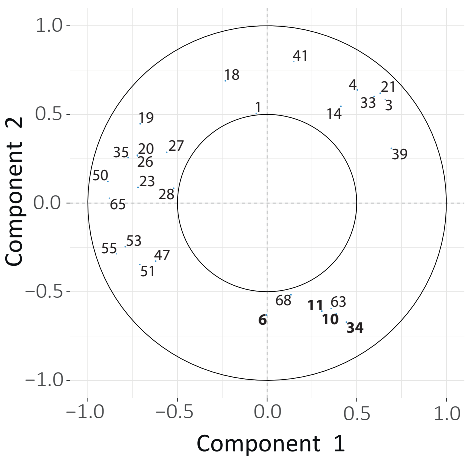


**Supplementary figure 2: PCA loadings**

A multilevel PCA was applied to individual samples, one sample being one participant at a specific timepoint, on the basis of cluster frequencies (scaled to unit variance). PCA loadings are demonstrated, with numbers refering to clusters identified in the unstimulated panel.

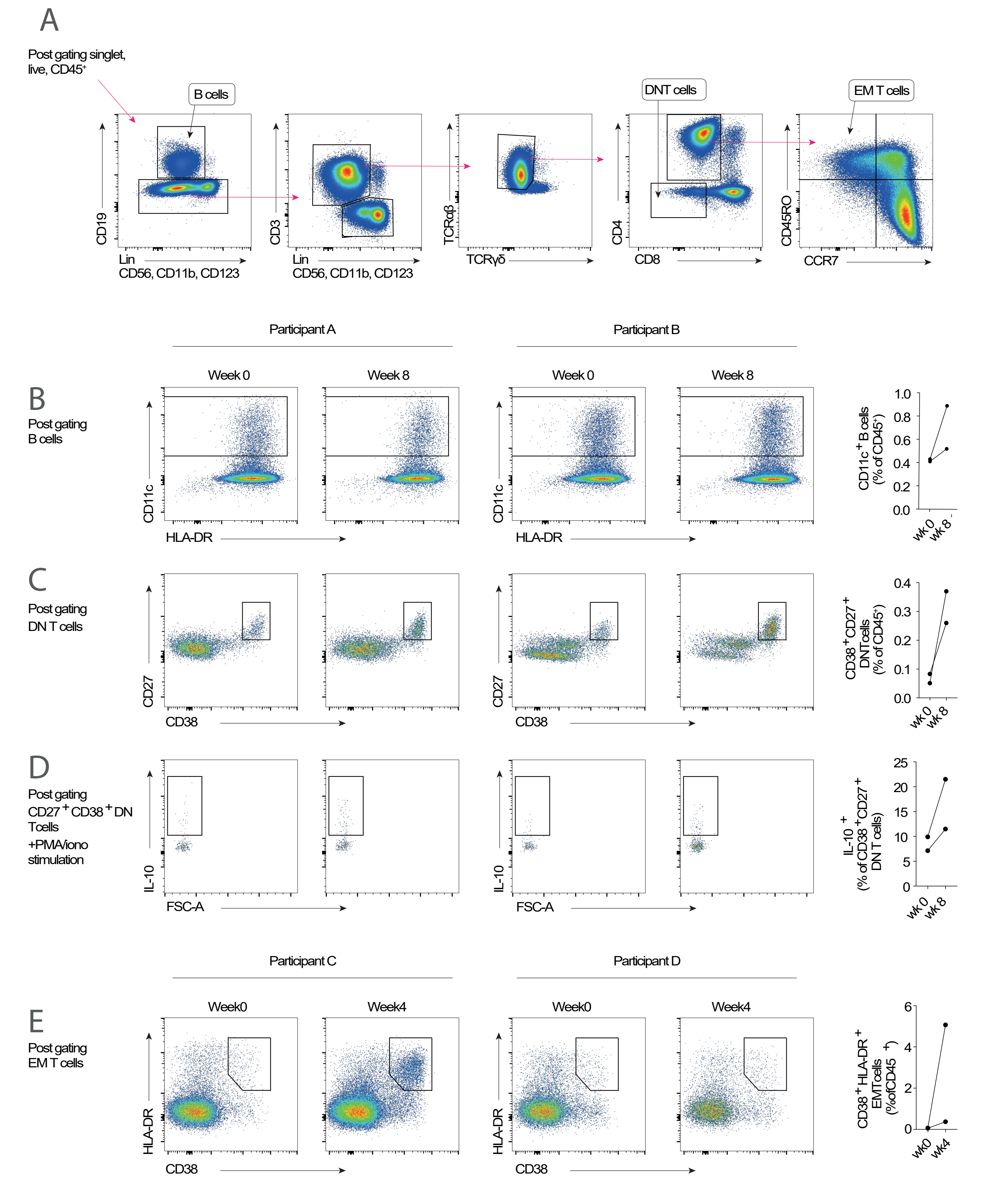


**Supplementary figure 3: Alterations in key lymphocyte populations confirmed via flow cytometry.**

A) Conventional flow cytometry was used to identify B cells, DN T cells, and EM T cells in two participants at week 0 and 4, and two participants at week 0 and 8 post controlled human *Schistosoma mansoni* infection. B, Increase in CD11c+ B cells at week 8 post infection. C) Increase in CD27+ CD38+ DN T cells at week at post infection. D) Expression of IL-10 in CD27+ CD38+ DN T cells post stimulation with PMA/ionomycin, increased at week 8 post infection. E, Increase in CD38+ HLA-DR+ EM T cells at week 4 post infection.

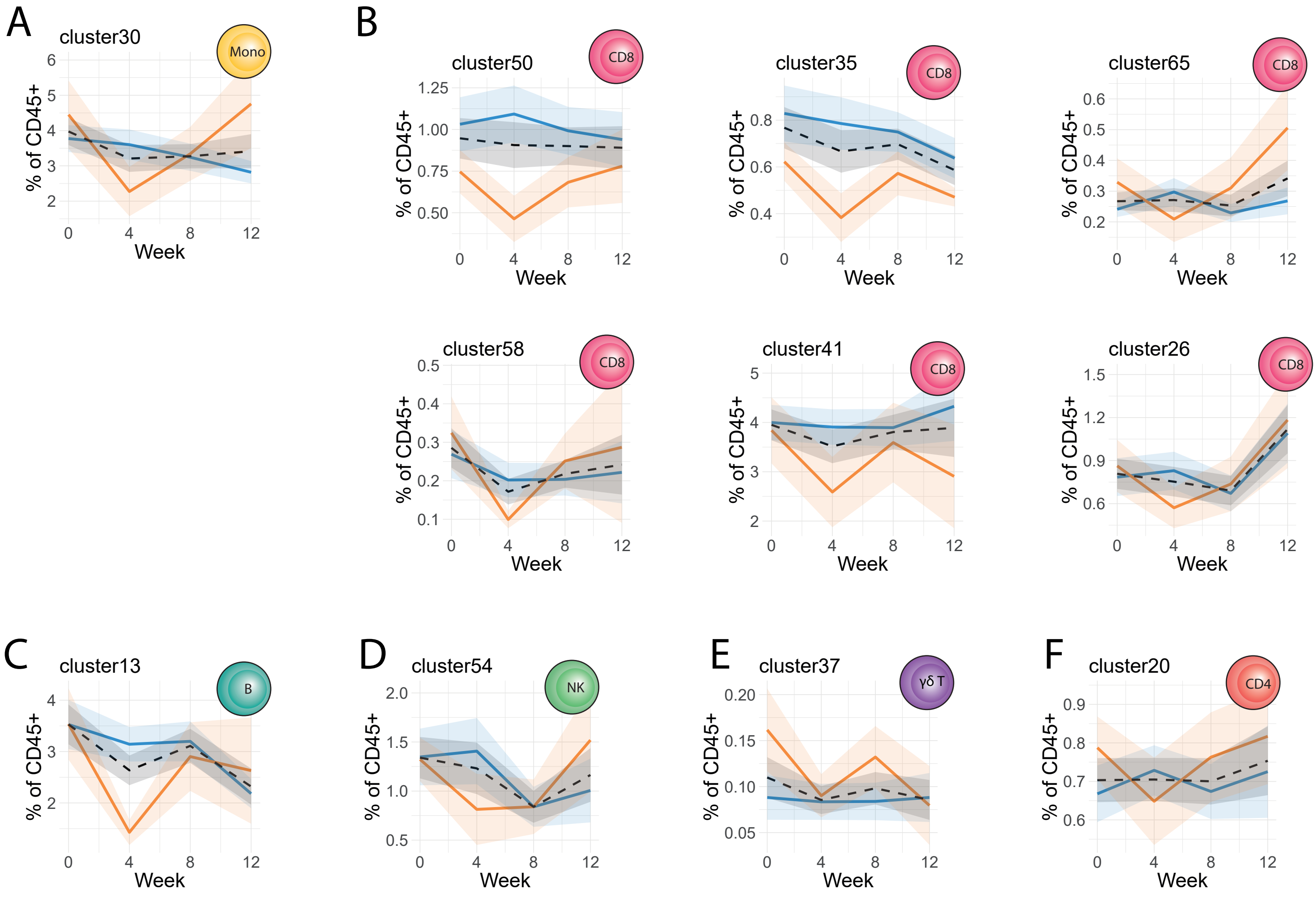


**Supplementary figure 4: Significantly altered cell clusters in unstimulated panel.** Ribbon plots showing frequency of labelled cluster over time. All colored ribbon plots show of cluster frequencies as percent of CD45+ cells. These are split and colored by symptoms, with lines representing the mean value and ribbon shading ± standard error of the mean (SEM). All analyses were performed on n=27 individual participants.

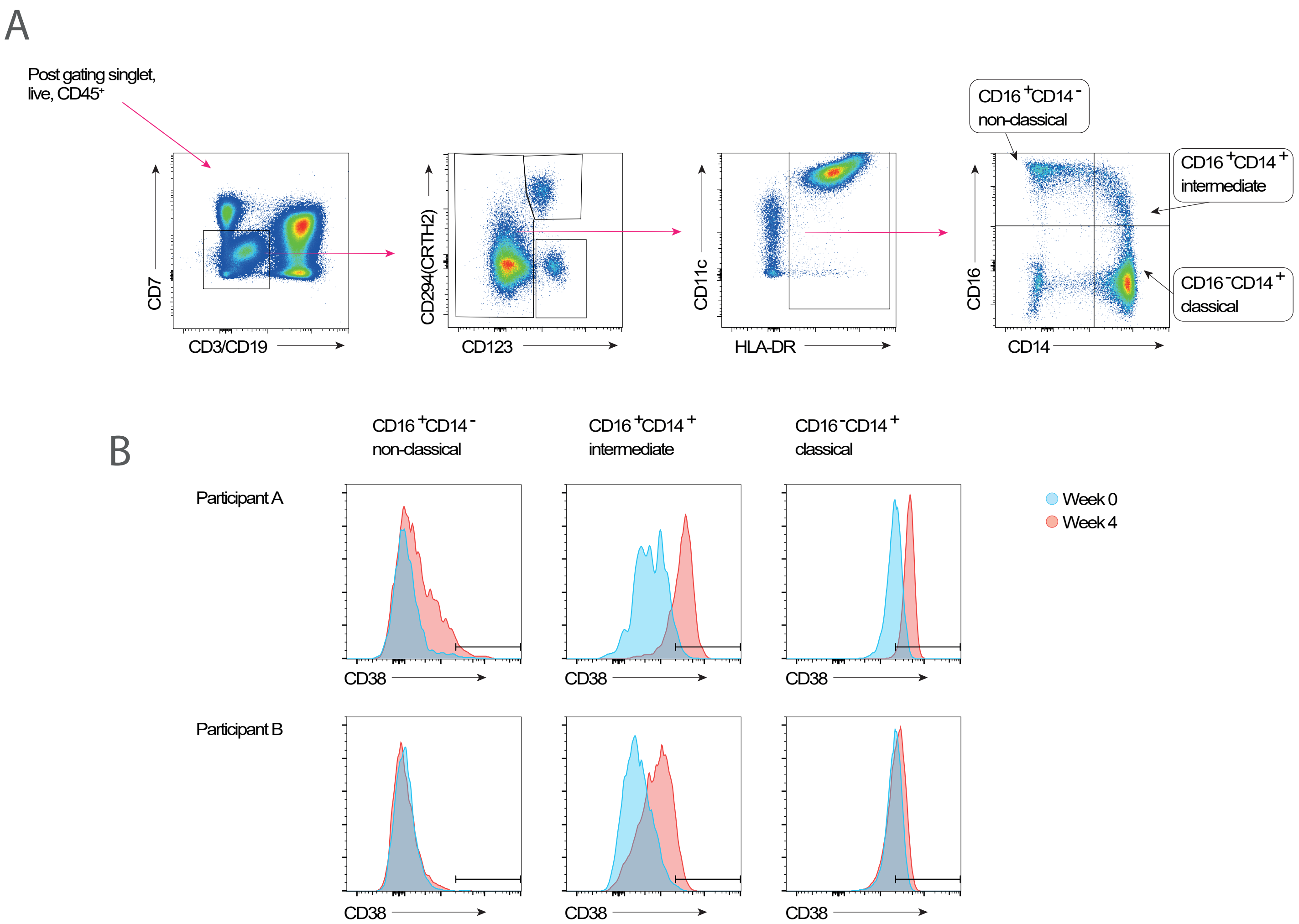


**Supplementary figure 5: Increase in monocyte CD38 expression observed with flow cytometry.**
A) Conventional flow cytometry was used to identify monocytes in two participants at week 0 and 4 post controlled human *Schistosoma mansoni* infection. B) CD38 expression in monocyte subsets over time.

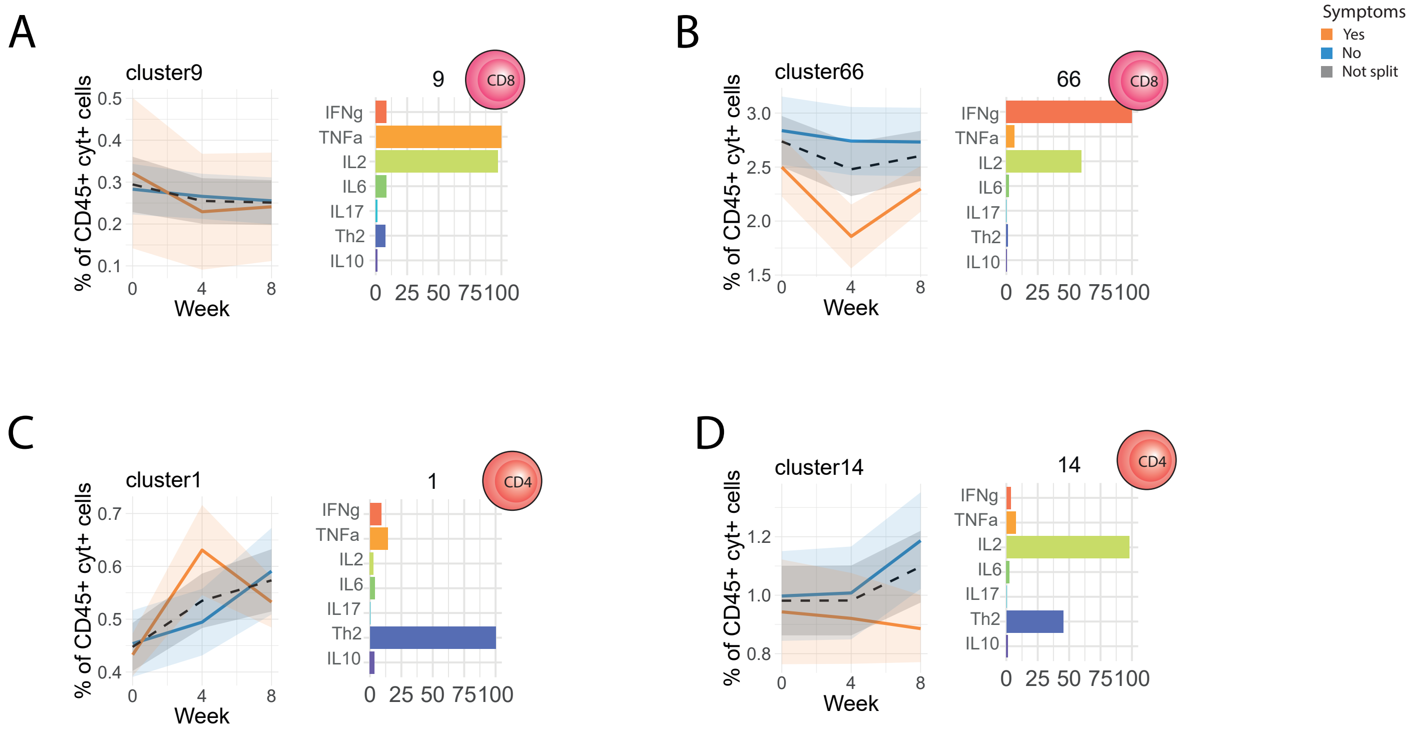


**Supplementary figure 6: Significantly altered cell clusters in stimulated panel.**

A-D) Colored ribbon plots show cluster frequencies as percent of cytokine+ CD45+ cells. These are split and colored by symptoms, with lines representing the mean value and ribbon shading ± standard error of the mean (SEM). Cytokine expression in each cluster is shown in the corresponding bar chart, colored by cytokine. All analyses were performed on n=27 individual participants.

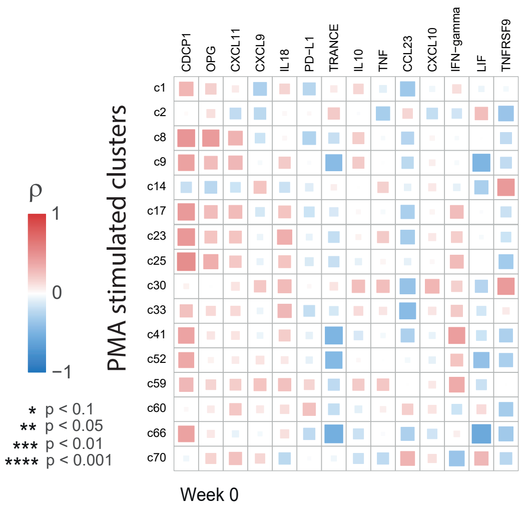


**Supplementary figure 7: Correlations between PMA stimulated cell clusters and serum cytokines at week 0.**

Correlation matrix comparing frequencies of cell clusters (rows) and serum cytokines (columns) at week 0. Color and size of squares are proportional to the spearmans ρ correlation coefficient. Cell clusters and serum cytokines selected to have significantly altered with time or symptoms, at a FDR<0.1. No significant correlations were found.

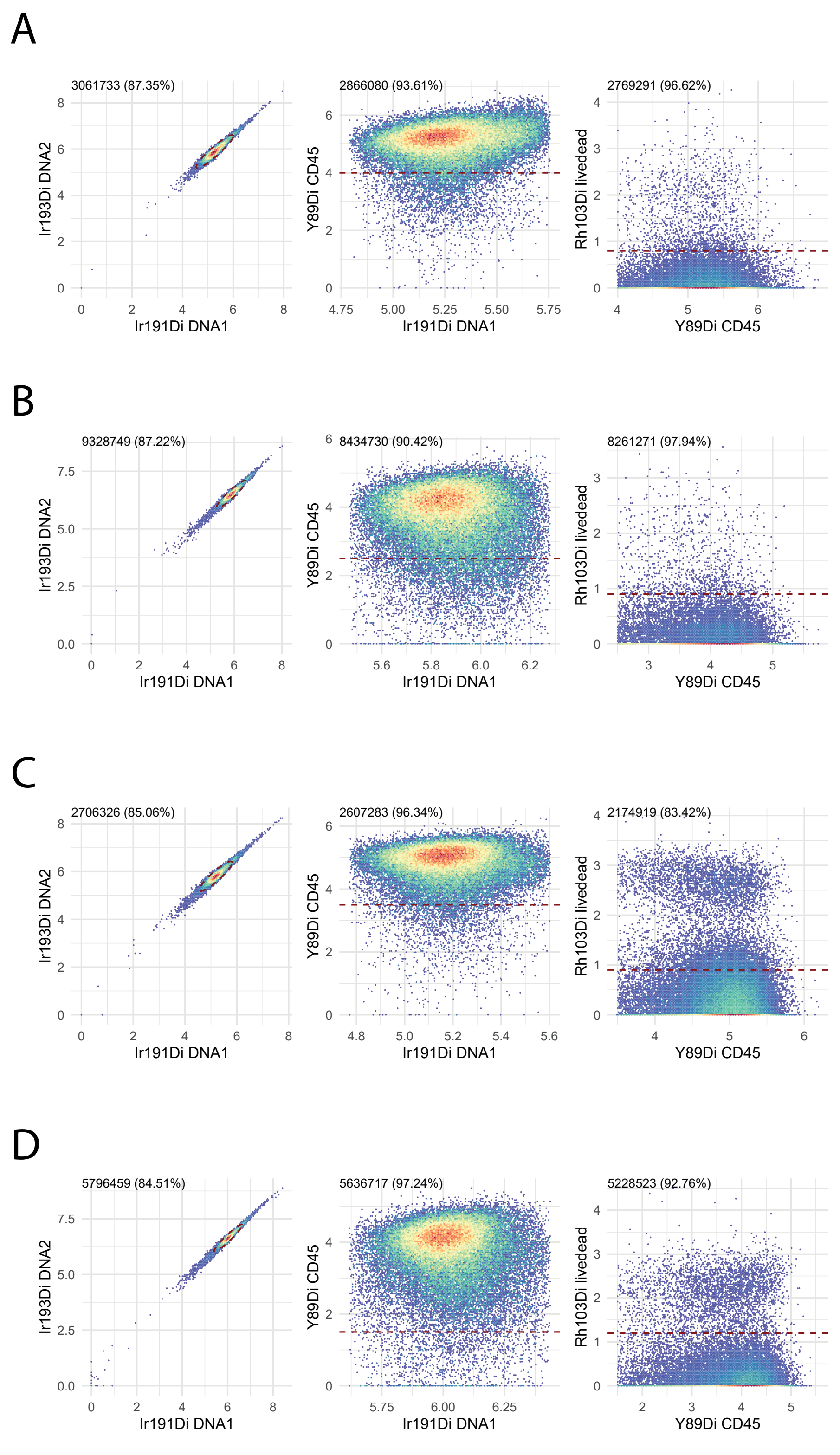


**Supplementary figure 8: Pre-gating of cellular data prior to unsupervised clustering.** Gating to select single, live, CD45+ cells for each batch, using the openCyto package in R. A representative example of one batch per pane is shown. A) Male cercariae infection unstimulated panel, B) Female cercariae infection unstimulated panel, C) Male cercariae infection stimulated panel, D) Female cercariae infection stimulated panel.

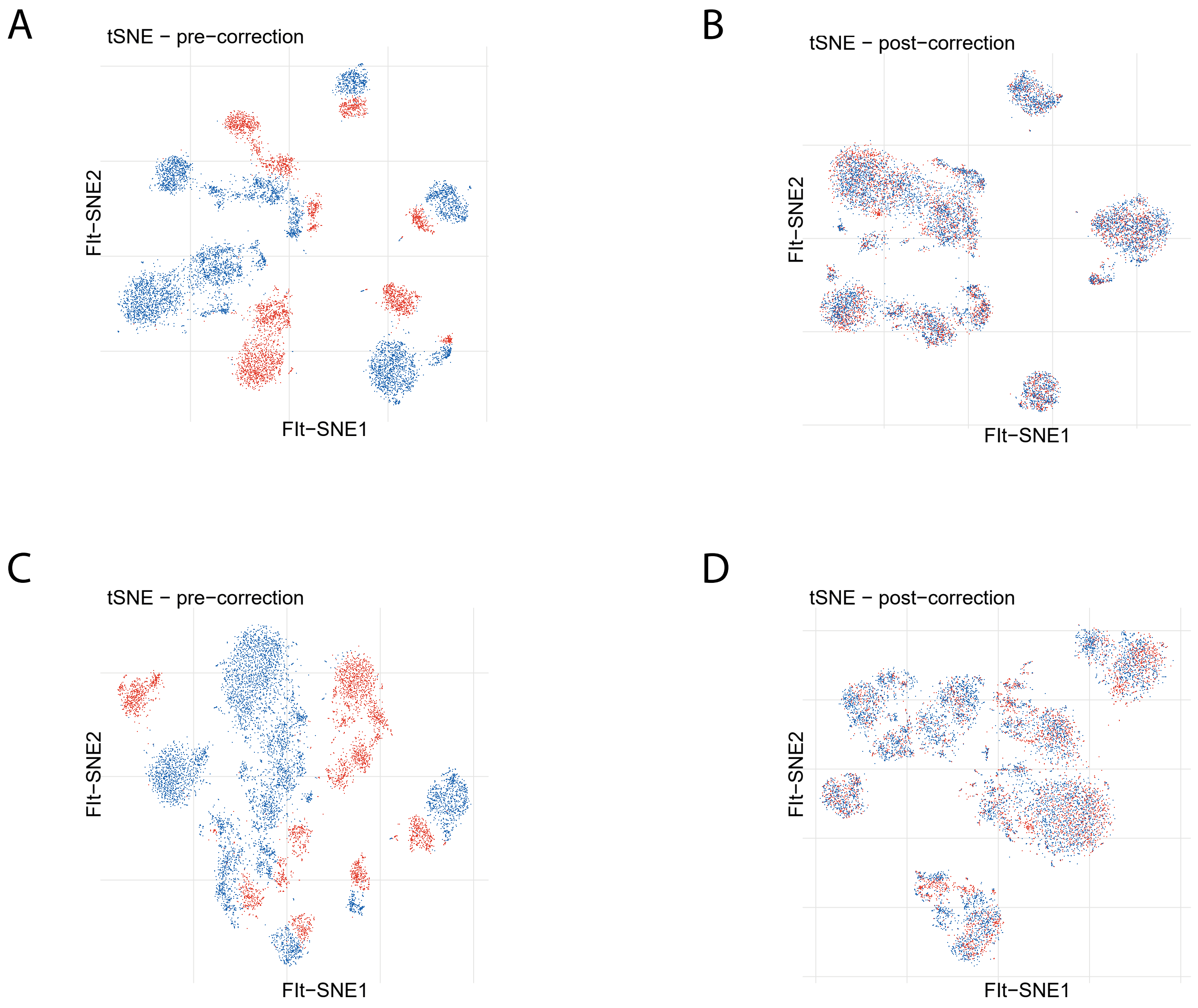


**Supplementary figure 9: Batch correction of male and female infection study.**
To correct technical variation between male and female infection studies, batch correction was performed using the CytoNorm packag. Pre (A&C) and post (B&D) batch correction tSNEs are shown, colored by study (red male infection, blue female infection). A&B show cells from the unstimulated panel. C&D show cells from the stimulated panel.
