## Supplementary tables for "Changes in pro inflammatory and regulatory immune responses during controlled human schistosome infection and the development of clinical symptoms"

| **Label** | **Specificity** | **Clone** | **Manufacturer** | **Cat no** | **Dilution** | | **Self conjugated** |
| --- | --- | --- | --- | --- | --- | --- | --- |
|  |  |  |  |  | **female** | **male** |  |
| ^89^Y | CD45 | HI30 | Fluidigm | 3089003B | 200 | 100 | No |
| ^115^In | CD278 (ICOS) | C398.4A | Biolegend | 313502 | 100 | N/A | Yes |
| ^115^In | CD45RA | HI100 | Fluidigm | 3155011B | N/A | 100 | No |
| ^141^Pr | CD196 (CCR6) | G034E3 | Fluidigm | 3141003A | 500 | 400 | No |
| ^142^Nd | CD19 | HIB19 | Fluidigm | 3142001B | 200 | 200 | No |
| ^143^Nd | CD117 (c-Kit) | 104D2 | Fluidigm | 3143001B | 500 | 100 | No |
| ^145^Nd | CD4 | RPA-T4 | Fluidigm | 3145001B | 200 | 200 | No |
| ^146^Nd | CD8a | RPA-T8 | Fluidigm | 3146001B | 500 | 200 | No |
| ^147^Sm | CD183 (CXCR3) | G025H7 | Biolegend | 353733 | 100 | 100 | Yes |
| ^148^Nd | CD14 | M5E2 | Biolegend | 301843 | 100 | 100 | Yes |
| ^149^Sm | CD25 (IL-2Ra) | 2A3 | Fluidigm | 3149010B | 500 | 100 | No |
| ^150^Nd | CD185 (CXCR5) | J252D4 | BioLegend | 356902 | 500 | 100 | Yes |
| ^151^Eu | CD123 | 6H6 | Fluidigm | 3151001B | 500 | 200 | No |
| ^152^Sm | TCRγδ | 11F2 | Fluidigm | 3152008B | 200 | 50 | No |
| ^153^Eu | CD7 | CD7-6B7 | Fluidigm | 3153014B | 100 | 200 | No |
| ^154^Sm | CD163 | GHI/61 | Fluidigm | 3154007B | 200 | 100 | No |
| ^155^Gd | CD45RA | HI100 | Fluidigm | 3155011B | 200 | N/A | No |
| ^155^Gd | CD278 (ICOS) | C398.4A | Biolegend | 313502 | N/A | 50 | No |
| ^156^Gd | CD294 (CRTH2) | BM16 | BioLegend | 350102 | 100 | 100 | Yes |
| ^158^Gd | CD122 (IL-2Rb) | TU27 | BioLegend | 339015 | 200 | 100 | Yes |
| ^159^Tb | CD197 (CCR7) | G043H7 | Fluidigm | 3159003A | 200 | 100 | No |
| ^161^Dy | KLRG1 (MAFA) | REA261 | Miltenyi | 130-126-458 | 200 | 200 | Yes |
| ^162^Dy | CD11c | Bu15 | Fluidigm | 3162005B | 100 | 100 | No |
| ^164^Dy | CD161 | HP-3G10 | Fluidigm | 3164009B | 200 | 100 | No |
| ^165^Ho | CD127 (IL-7Ra) | AO19D5 | Fluidigm | 3165008B | 500 | 200 | No |
| ^167^Er | CD27 | O323 | Fluidigm | 3167002B | 200 | 200 | No |
| ^168^Er | HLA-DR | L243 | Biolegend | 307651 | 200 | 200 | Yes |
| ^170^Er | CD3 | UCHT1 | Fluidigm | 317001B | 150 | 100 | No |
| ^171^Yb | CD28 | CD28.2 | Biolegend | 302902 | 500 | 100 | Yes |
| ^172^Yb | CD38 | HIT2 | Fluidigm | 3172007B | 200 | 200 | No |
| ^173^Yb | CD45RO | UCHL1 | BioLegend | 304239 | 200 | 100 | Yes |
| ^174^Yb | CD335 (NKp46) | 92E | BioLegend | 331902 | 500 | 100 | Yes |
| ^175^Lu | CD279 (PD-1) | EH12.2H7 | Fluidigm | 3175008B | 500 | 100 | No |
| ^176^Yb | CD56 | NCAM16.2 | Fluidigm | 3176008B | 500 | 200 | No |
| ^209^BI | CD16 | 3G8 | Fluidigm | 3209002B | 200 | 200 | No |
| ^169^Tm | GATA3 | REA174 | Miltenyi | 130-108-061 | 100 | 50 | Yes |
| ^166^Er | Tbet | 4B10 | Biolegend | 644802 | 100 | 100 | Yes |
| ^163^Dy | CD152 (CTLA-4) | BNI3 | Biolegend | 369602 | 200 | 100 | Yes |
| ^160^Gd | FOXp3 | PCH101 | Fluidigm | 3162011A | 100 | 50 | Yes |

**Supplementary Table 1: Antibodies used for mass cytometry, unstimulated panel.**

| **Label** | **Specificity** | **Clone** | **Manufacturer** | **Cat no** | **Dilution** | | **Self conjugated** |
| --- | --- | --- | --- | --- | --- | --- | --- |
|  |  |  |  |  | **female** | **male** |  |
| ^89^Y | CD45 | HI30 | Fluidigm | 3089003B | 200 | 100 | No |
| ^115^In | CD45RA | HI100 | BioLegend | 304102 | 100 | 100 | Yes |
| ^141^Pr | CD196 (CCR6) | G034E3 | Fluidigm | 3141003A | 500 | 400 | No |
| ^142^Nd | CD19 | HIB19 | Fluidigm | 3142001B | 200 | 200 | No |
| ^143^Nd | CD117 (c-Kit) | 104D2 | Fluidigm | 3143001B | 500 | 100 | No |
| ^145^Nd | CD4 | RPA-T4 | Fluidigm | 3145001B | 100 | 200 | No |
| ^146^Nd | CD8a | RPA-T8 | Fluidigm | 3146001B | 500 | 200 | No |
| ^147^Sm | CD183 (CXCR3) | G025H7 | Biolegend | 353733 | 100 | 100 | Yes |
| ^148^Nd | CD14 | M5E2 | Biolegend | 301843 | 200 | 100 | Yes |
| ^149^Sm | CD25 (IL-2Ra) | 2A3 | Fluidigm | 3149010B | 500 | 100 | No |
| ^150^Nd | CD185 (CXCR5) | J252D4 | BioLegend | 356902 | 200 | 100 | Yes |
| ^151^Eu | CD123 | 6H6 | Fluidigm | 3151001B | 500 | 200 | No |
| ^152^Sm | TCRγδ | 11F2 | Fluidigm | 3152008B | 200 | 50 | No |
| ^153^Eu | CD7 | CD7-6B7 | Fluidigm | 3153014B | 200 | 200 | No |
| ^156^Gd | CD294 (CRTH2) | BM16 | BioLegend | 350102 | 100 | 100 | Yes |
| ^158^Gd | CD122 (IL-2Rb) | TU27 | BioLegend | 339015 | 200 | 100 | Yes |
| ^159^Tb | CD197 (CCR7) | G043H7 | Fluidigm | 3159003A | 100 | 100 | No |
| ^161^Dy | KLRG1 (MAFA) | REA261 | Miltenyi | 130-126-458 | 200 | 200 | Yes |
| ^162^Dy | CD11c | Bu15 | Fluidigm | 3162005B | 200 | 100 | No |
| ^164^Dy | CD161 | HP-3G10 | Fluidigm | 3164009B | 200 | 100 | No |
| ^165^Ho | CD127 (IL-7Ra) | AO19D5 | Fluidigm | 3165008B | 500 | 200 | No |
| ^167^Er | CD27 | O323 | Fluidigm | 3167002B | 200 | 200 | No |
| ^168^Er | HLA-DR | L243 | Biolegend | 307651 | 200 | 200 | Yes |
| ^170^Er | CD3 | UCHT1 | Fluidigm | 317001B | 150 | 100 | No |
| ^171^Yb | CD28 | CD28.2 | Biolegend | 302902 | 200 | 100 | Yes |
| ^172^Yb | CD38 | HIT2 | Fluidigm | 3172007B | 100 | 200 | No |
| ^173^Yb | CD45RO | UCHL1 | BioLegend | 304239 | 200 | 100 | Yes |
| ^175^Lu | CD279 (PD-1) | EH12.2H7 | Fluidigm | 3175008B | 500 | 100 | No |
| ^176^Yb | CD56 | NCAM16.2 | Fluidigm | 3176008B | 500 | 200 | No |
| ^144^Nd | IL-2 | MQ117H12 | Fluidigm | 3144021B | 200 | 400 | No |
| ^155^Gd | IFNg | B27 | BioLegend | 506521 | 200 | 400 | Yes |
| ^160^Gd | TNFa | MAb11 | BioLegend | 502941 | 200 | 400 | Yes |
| ^166^Er | IL-10 | JES39D7 | Fluidigm | 3166008B | 200 | 400 | No |
| ^169^Tm | IL-4 | MP4-25D2 | Biolegend | 500802 | 200 | 400 | Yes |
| ^169^Tm | IL-5 | TRFK5 | BioLegend | 504302 | 200 | 400 | Yes |
| ^169^Tm | IL-13 | JES105A2 | Biolegend | 501902 | 200 | 400 | Yes |
| ^163^Dy | IL-17 | BL168 | Biolegend | 512302 | 500 | 400 | Yes |
| ^154^Sm | IL-6 | MQ2-13A5 | Biolegend | 501102 | 100 | 400 | Yes |

**Supplementary Table 2: Antibodies used for mass cytometry, PMA/ionomycin stimulated panel.**
